# The Sparse Matrix of Drug Discovery: Sex, Race, and a Genomic Equity Index Across 40.8 Million Patients in 77,770 Clinical Trials

**DOI:** 10.64898/2026.05.14.26353197

**Authors:** Anil Bajnath, Allana Roach, Rajini Haraksingh, Irman Forghani, Alexander N. Evans, Elena Cyrus, Dexter Hadley

**Affiliations:** American Board of Precision Medicine (ABOPM), Florida, USA; CANONIC Foundation, Orlando, FL, USA; American Cancer Society, Atlanta, GA, USA; The University of the West Indies, St. Augustine, Trinidad & Tobago; Yale-TCC Consortium, New Haven, CT, USA; Mount Sinai Medical Center of Florida, Miami Beach, FL, USA; Howard University College of Medicine and Howard University Hospital, Washington, DC, USA; University of Central Florida College of Medicine, Orlando, FL, USA

## Abstract

Demographic representation in clinical trials and downstream genomic databases has been documented to be skewed across two decades, but the structural origin of the skew remains contested. Here we test whether a single upstream protocol-writing bias drives demographic skew on both axes (race and sex) at the population scale of clinical-trial enrollment, and whether the same skew compounds at the genomic-database layer that informs precision-medicine inference.

The hypothesis is anchored to a previously characterized clinical-care bias. The senior author has previously shown that the named sex disparity in coronary artery bypass grafting (CABG) does not originate at the operating-room door but in diagnostic timing, with women experiencing significantly longer first-encounter to diagnostic-catheterization intervals than men, while no sex difference appears in cath to CABG timing^9^. The bedside record shows what looks like sex-biased surgery; the upstream record reveals a bias set in who is investigated for cardiac disease at all. CABG is the bedside template; clinicaltrial enrollment is the population-scale analogue.

In the trial record, the CABG signature reproduces, on both demographic axes. On the race axis, African-derived enrollment declines Phase I → Phase III monotonically across all sponsor classes; the industry × academia joint trial shows the steepest gradient of all (Phase III 8.2%; Δ Phase I → III = +4.65 pp; OR = 1.62; *P* < 10^−100^), with academic execution under industry-written protocols laundering catchment-area diversity out. On the sex axis, female enrollment runs the opposite phase direction (Phase I 41.4% → Phase III 48.7% in the eligibility-ALL subset, n = 37,737), with the inversion concentrated in industry-solo trials (+10.0 pp); 99.3% of trials report sex against 61.6% reporting race. The two axes are statistically independent at the trial level (joint CABG-bias quadrant occupancy 24.81% vs 24.76% expected under independence; χ^2^ = 0.13, *P* = 0.72; Pearson r = 0.016), one upstream cause with two parallel readouts, not one reinforcing race × sex effect.

Extended to the genomic-database layer, the pattern compounds. The GEI ranks European-derived populations at 0.98; eight others score 0.58 or below; the global equity gap is 3.94 billion people receiving precision medicine calibrated for the 11% of humanity who are European-derived. The race-axis primary-data recomputation of GWAS Catalog ancestry share yields 88.3% European-derived participation, not the published 78% (Mills & Rahal 2020), the gap has widened, not narrowed. Top and bottom GEI rankings are robust in 100% of 10,000 Dirichlet weight permutations. The same architecture is portable to a parallel Sex Equity Index for the orthogonal axis.

The hypothesis is supported at every layer we measured. The structural locus across all three, bedside, trial, genomic database, is one upstream governance function. Boards and credentialing bodies that govern protocol design, independent of the sponsor, are the structural intervention point at every layer.

## Introduction

Three separate scholarly traditions have measured what is, on inspection, the same structural failure at three points along the drug-development pipeline. The clinical-care literature documents named demographic biases at the bedside^9^. The clinical-trial literature documents enrollment skew on race and sex^1–6^. The genomic-database literature documents downstream undersampling on ancestry^7,10–12^. Each tradition treats its layer as the locus. None has connected them. Here we argue that all three measure the same upstream protocol-writing function, and we test that argument by analyzing 77,770 ClinicalTrials.gov studies (40.8 million participants, data freeze 1 May 2026) jointly on race and sex, anchored at the named bedside phenomenon for which the upstream-bias hypothesis is best characterized: coronary artery bypass grafting.

### An upstream bias visible at the bedside: CABG

Coronary artery bypass grafting is a textbook case of a clinical-care bias whose locus sits one step earlier than the decision visible in the surgical record. The senior author has previously shown that the sex disparity in CABG does not originate at the operating-room door^9^. In an analysis of approximately 32,000 patients with coronary artery disease, women experienced a significantly longer interval between their first physician encounter indicative of CAD and their first diagnostic cardiac catheterization than men, while the interval from diagnostic catheterization to CABG itself showed no sex difference^9^. The bias is in who is investigated for cardiac disease at all, not in who is offered the procedure once disease is established. The downstream record, the bypass operation, its outcomes, the women whose surgeries were delayed or never happened, looks like sex-biased surgery. The upstream record, who entered the diagnostic pipeline, is where the bias lives.

CABG matters here as a conceptual template, not as a sex-only story. It names a pattern: a recorded clinical-care decision whose demographic skew is set by an upstream investigation step. The same pattern, if it exists in the trial pipeline, would predict that demographic skew in Phase III labels and post-market evidence is set in Phase I/II protocol-writing, before any efficacy data exists.

### From bedside to protocol: trial enrollment is the population-scale upstream step

Clinical-trial enrollment is the population-scale analogue of the bedside investigation step. Every Phase I → Phase III enrollment decision is made before the trial-result record exists, in the same upstream way the diagnostic-investigation decision precedes the surgical record. If the CABG-style upstream-bias model generalizes, the ClinicalTrials.gov enrollment record should carry the same demographic-skew signatures the bedside literature documents, visible both as the racial-enrollment bias the trial-disparity literature has documented for two decades^2–4^, and as a parallel sex-enrollment skew on the same trials. We test both, on the same 77,770-trial corpus, and we test their joint distribution.

## Results

### Dataset and reporting compliance

#### Reporting compliance: where the data is and where it is not

The two demographic axes have asymmetric data shadows. Of 77,770 studies with posted results, 47,889 (61.6%) reported race or ethnicity for their participants, representing 40,791,212 individuals across two decades (2005–2026). Sex, by contrast, was reported by 77,240 trials (99.3%). The data on the sex axis has been there; the analytical attention has not. The race-axis sparse matrix that follows is necessarily computed on the 61.6% of trials that report race; the sex-axis findings later in the paper are computed on the 99.3% that report sex (filtered to the n = 37,737 eligibility-ALL subset where the trial protocol did not pre-restrict enrollment to a single sex; Methods). Where this paper reports a quantity for one axis but not the other, the underlying compliance asymmetry is the reason, not an analytical choice.

### Race-axis findings

#### The sparse matrix: race-axis evidence deserts

We extracted baseline race from the ClinicalTrials.gov v2 `resultsSect ion.baselineCharacteristicsModule`, normalized to the NIH/OMB standard, and grouped therapeutic areas by MeSH condition code into 15 categories (Methods). For each therapeutic area × population pair we computed an Evidence Score:

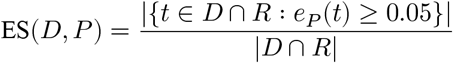

where D is the set of trials in a therapeutic area, R is the set of trials with race data, and *e*_*P*_ (*t*) is the fractional enrollment of population P in trial t. We classified cells as ADEQUATE (ES ≥ 0.30), WEAK (0.10–0.30), INSUFFICIENT (0.01–0.10) or ZERO (<0.01).

The result is shown in **Fig. 1**. The European-derived column is uniformly ADEQUATE across all 15 therapeutic areas (range 75.8–95.5%). The African-derived column is ADEQUATE everywhere but with substantial range (39.1–74.7%). The Asian-derived column oscillates between ADEQUATE and WEAK. The Indigenous American column is INSUFFICIENT (1.9–5.7%) across 15 of 15 therapeutic areas. The Pacific Islander column is ZERO or INSUFFICIENT across 15 of 15. **Indigenous American and Pacific Islander populations occupy evidence deserts in 100% of therapeutic areas: there is no drug class, not one, for which either population has ADEQUATE clinical trial evidence**. The aggregate is a governance failure that conceals the therapeutic-area-specific disparities which determine whether a patient receives evidence-based treatment.

**Fig. 1.**
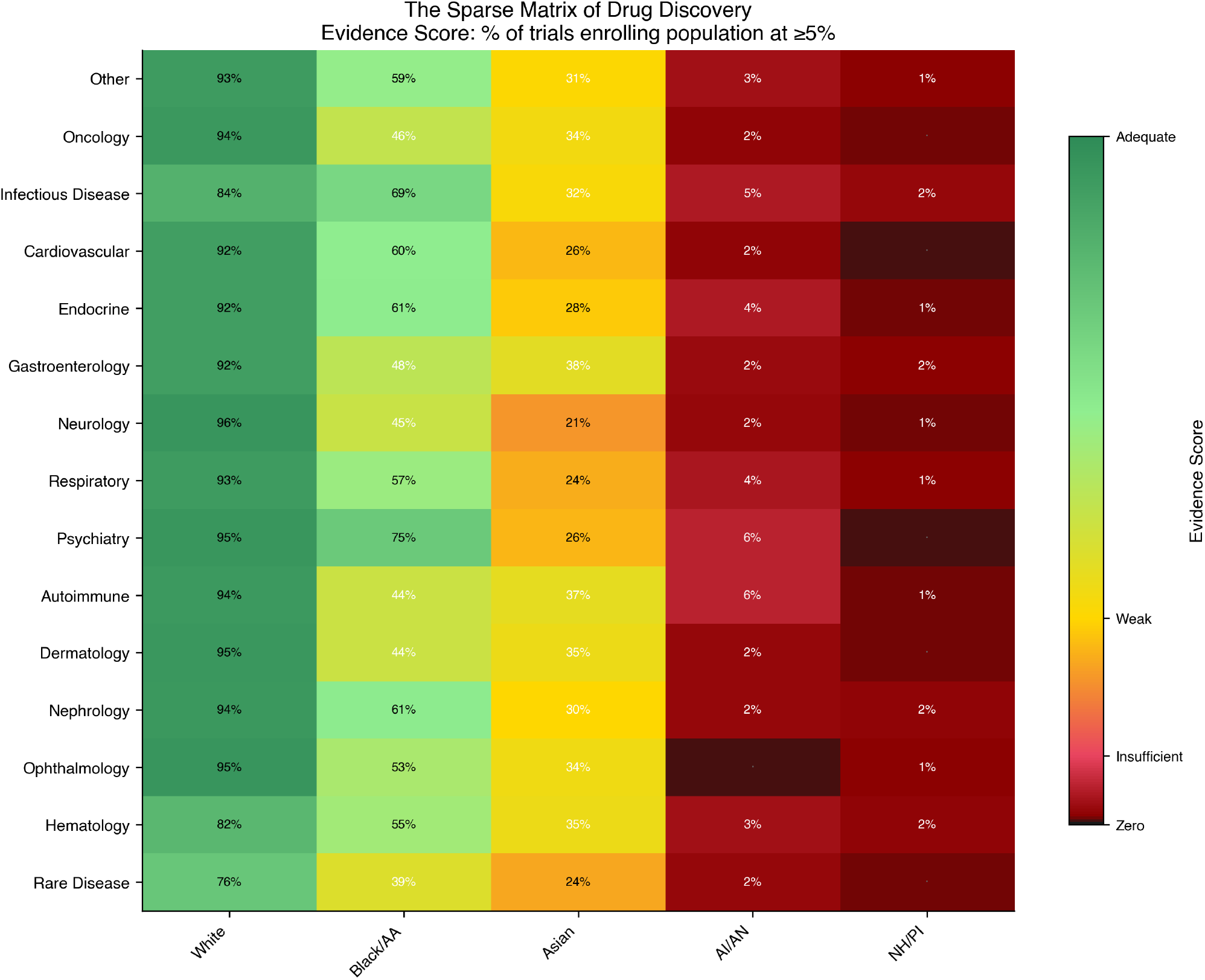
The Sparse Matrix of Drug Discovery. Evidence scores across 15 therapeutic areas × 5 ancestry populations. Cells colored by Evidence Score (ES = proportion of trials in the therapeutic area enrolling that population at ≥5%) and classified ADEQUATE (ES ≥ 0.30), WEAK (0.10–0.30), INSUFFICIENT (0.01–0.10), or ZERO (<0.01). The European-derived column is ADEQUATE across all 15 therapeutic areas. Indigenous American and Pacific Islander populations occupy evidence deserts across all 15 therapeutic areas, no drug class has adequate trial evidence for either population. N = 47,889 trials with race data (61.6% of 77,770 total studies with posted results), 40,791,212 participants over two decades (2005–2026; data freeze 1 May 2026).

Every therapeutic area failed Census proportionality (all χ^2^ > 3,000; all *P* < 10^−10^; all surviving Bonferroni correction at α = 0.003 for 15 comparisons). The headline aggregate, 18.4% African-derived enrollment exceeding the 13.4% Census share, is the aggregate deception: it is driven almost entirely by infectious disease trials (HIV, COVID-19) that enrolled African-derived participants at 26.9%. Oncology is 8.8%. Neurology is 9.8%. Dermatology is 9.4%. African-derived men carry the highest cancer mortality of any racial group in the United States^8^ yet enroll in oncology trials at 59% of their disease burden.

#### The phase funnel on both axes

The sparse matrix is a snapshot. The pipeline is a temporal process. The CABG model predicts that demographic skew in the downstream record (Phase III labels, post-market evidence) is set in the upstream investigation step (Phase I/II protocol design). We test that prediction by stratifying enrollment by trial phase on both demographic axes (**Fig. 2**).

**Fig. 2.**
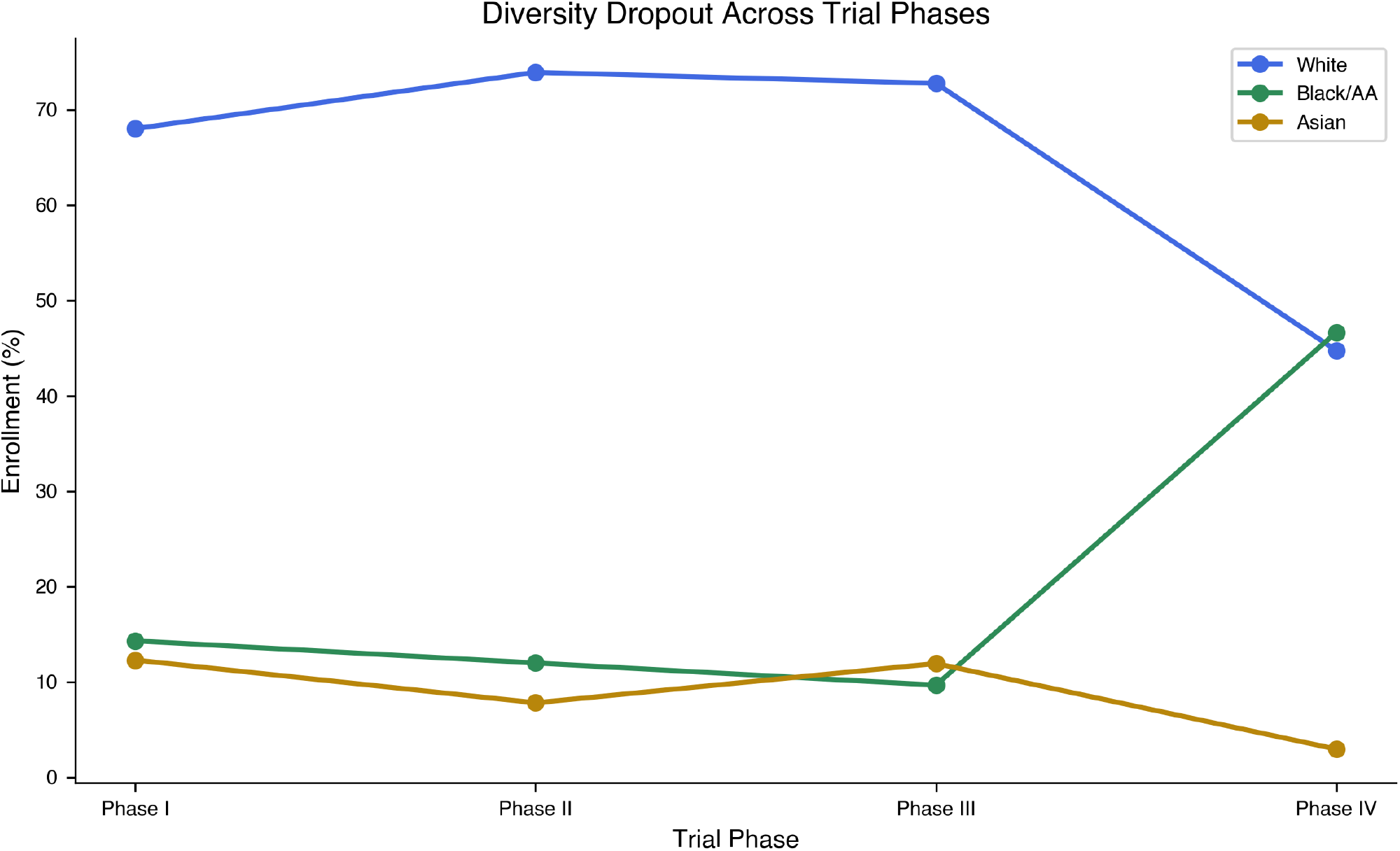
The phase funnel on both demographic axes. (a) African-derived enrollment from Phase I to Phase III, stratified by industry vs NIH vs federal sponsor. African-derived enrollment declines monotonically across all sponsor types, with the industry slope 3.3× steeper than NIH (−3.1 vs −0.95 pp/phase). In industry oncology Phase III, African-derived enrollment is 1.8% against a cancer mortality burden of 15%. (b) Female enrollment from Phase I to Phase III in the eligibility-ALL subset (n = 37,737). The sex gradient runs in the opposite direction: Phase I 41.4% → Phase III 48.7%, with the female deficit concentrated in the early-phase trials where pharmacokinetic baselines are set. Both gradients are visible on the same trials, originating upstream of the recorded efficacy result. The risk (race) and the absence (sex) both flow in early; the evidence flows out late.

##### The race axis

African-derived enrollment declines from Phase I to Phase III in both industry-sponsored and publicly-funded trials, but the decline is much sharper in industry (**Table 1**).

**Table 1.** Phase I → III African-derived enrollment by sponsor type.

| Sponsor | Phase I | Phase II | Phase III | Slope (pp/phase) |
| --- | --- | --- | --- | --- |
| Industry | 14.3% | 10.2% | 8.1% | −3.1 |
| NIH | 22.4% | 21.1% | 20.5% | −0.95 |
| Federal | 23.8% | 22.7% | 22.1% | −0.85 |

Industry slope 3.3× steeper than NIH; industry enrolls African-derived patients at 2.5× lower rates than NIH in Phase III (χ^2^ = 73,408, *P* < 2.2 × 10^−16^).

The pooled slope across sponsors is −2.3 percentage points per phase (linear regression on phase ordinal, *P* = 0.0004). African-derived patients are 47% more likely to be enrolled in industry safety trials than in the efficacy trials that determine FDA labels (odds ratio Phase I vs Phase III = 1.30; Fisher’s exact *P* < 2.2 × 10^−16^). The pattern reaches its extreme in industry oncology: Phase I 4.5% African-derived, Phase II 4.1%, Phase III 1.8% (OR Phase I vs Phase III = 2.53; Fisher’s exact *P* < 2.2 × 10^−16^). Cancer mortality parity would predict 15% African-derived enrollment; at parity, **83**,**614 African-derived patients are missing** from industry oncology Phase III trials. The risk flows in. The evidence flows out.

The **sex axis** runs in the opposite direction on the same trials. In the n = 37,737 eligibility-ALL analytic subset (where the protocol did not pre-restrict to a single sex), Phase I averages 41.4% female and Phase III averages 48.7% female. Female enrollment is lowest in the early phases that calibrate pharmacokinetic and pharmacodynamic baselines, highest in the efficacy phase that produces the FDA label, the inverse of the African-derived gradient. The female deficit is concentrated in exactly the phases where African-derived enrollment is highest, and vice versa.

Two demographic axes, two opposite-direction Phase I → Phase III gradients, both visible on the same 77,770 trials, both originating in the same upstream protocol-writing step. This is the CABG signature at population scale: demographic skew set upstream of the recorded decision, not in the decision itself. Whether the upstream skew is the same failure mode on both axes (i.e. whether the trials that exclude non-European-derived populations are also the trials that exclude women) is the joint-distribution question we resolve in **§The two axes are independent failure modes** below.

#### Named sponsors

We identified six pharmaceutical sponsors whose Phase III trials exceeded 80,000 participants with Phase III African-derived enrollment below 3%. Together, these sponsors contributed over 670,000 Phase III participants, a cohort larger than most national biobanks, with effectively zero African-derived evidence. In every case, Phase I enrollment was substantially higher than Phase III, indicating systematic transition out of the efficacy pipeline for the populations that bore the safety burden.

| Sponsor | Phase III N | Phase III<br>African-derived | Phase I<br>African-derived | $\Delta$ (I – III) |
| --- | --- | --- | --- | --- |
| Novo Nordisk | 163,297 | 4.7% | 50.8% | +46.1 pp |
| GlaxoSmithKline | 117,792 | 7.0% | 33.1% | +26.2 pp |
| Eli Lilly | 333,687 | 5.1% | 14.0% | +8.9 pp |
| Amgen | 153,183 | 2.9% | 11.6% | +8.7 pp |
| AstraZeneca | 155,638 | 4.9% | 13.5% | +8.5 pp |
| Merck | 311,536 | 5.1% | 10.6% | +5.5 pp |

Spearman correlation of industry Phase III African-derived enrollment over 2010–2025: ρ = 0.459, *P* = 0.074 (n.s.). Fifteen years of public diversity commitments produced no measurable change in the underlying enrollment patterns.

#### The sponsor-class gradient: who actually writes the protocol

The lead-sponsor field is one structural variable; the joint-trial structure is another, and it predicts demographics more sharply on both axes. We extracted the full `sponsorColl aboratorsModule` (lead sponsor, collaborators, responsible party) for all 77,770 trials and bucketed each into one of four mutually exclusive structures (industry-led, industry × academia joint, academia-led, government-led), then ran the Phase I → III gradient for each on both demographic axes (**Fig. 3, Table 2**).

**Table 2.** Phase I → Phase III enrollment by sponsor structure on both demographic axes. Race-axis Δ in pp of African-derived enrollment; sex-axis Δ in pp of female enrollment, restricted to eligibility-ALL trials.

| Bucket | Trials | Phase III N | Black% | Race $\Delta$ (I–III) | Race OR /<br>$P$ | Female $\Delta$<br>(I→III) |
| --- | --- | --- | --- | --- | --- | --- |
| Industry-led | 34,302 | 6,110,912 | 7.6% | +1.95 pp | 1.27 /<br><10<sup>−100</sup> | +10.0 pp |

| Bucket | Trials | Phase III N | Black% | Race $\Delta$ (I–III) | Race OR / $P$ | Female $\Delta$ (I→III) |
| --- | --- | --- | --- | --- | --- | --- |
| Industry × academia joint | 8,942 | 526,196 | **8.2%** | **+4.65 pp** | **1.62 / $<10^{-100}$** | +2.2 pp |
| Academia-led | 20,104 | 166,601 | 19.0% | +0.63 pp | 1.04 / n.s. | +0.1 pp |
| Government-led | 13,785 | 860,969 | 19.9% | +0.62 pp | 1.04 / n.s. | −0.7 pp |

**Fig. 3.**
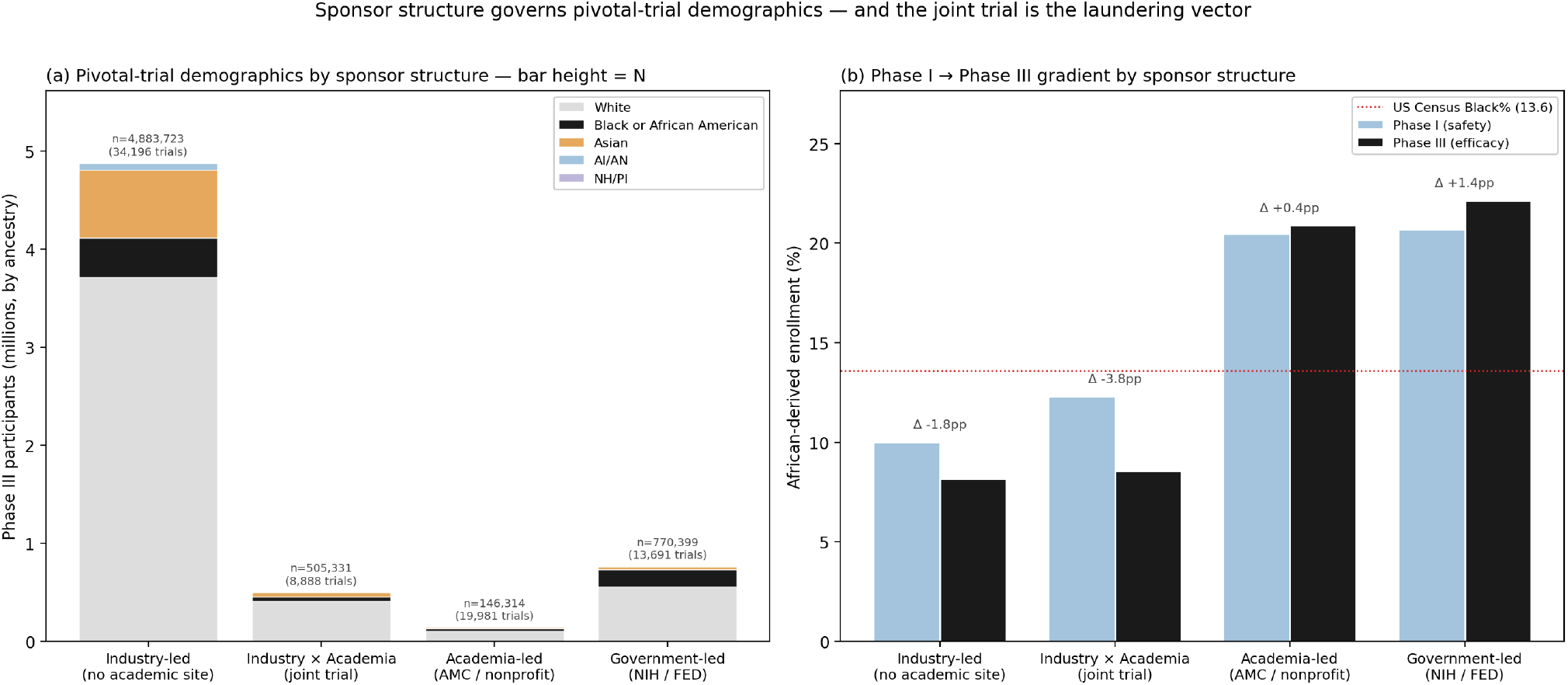
The sponsor-class gradient on both demographic axes. (a) Pivotal-trial demographics by sponsor structure across 77,770 trials, rendered as count-stacked bars with bar height = Phase III participant N. The industry-led bar (n = 6,110,912; 34,302 trials) is ∼37-fold taller than the academia-led bar (n = 166,601; 20,104 trials) and is dominantly white (76% White; 7.6% African-derived); the academia-led bar is more diverse (69% White; 19.0% African-derived) but represents an order-of-magnitude fewer participants. The largest pipeline is also the most homogeneous. (b) Phase I → Phase III African-derived gradient by bucket. The industry × academia joint trial shows the steepest extraction gradient (+4.65 pp; OR = 1.62; *P* < 10^−100^), more than twice the slope of industry-led trials alone. Academia-led and government-led trials show no meaningful race gradient (<1 pp). The protocol governs the demographics of the trial; the site does not. Panel-(c) female-share gradient analysis is reported in **Table 2** and the corresponding text.

The two axes tell the same structural story from different directions.

The **race axis** reproduces the established sponsor pattern. Academia-led and government-led trials show no meaningful extraction gradient, Phase I → Phase III African-derived enrollment differs by less than 1 percentage point in both buckets, and the catchment-area populations of academic medical centers and the recruitment networks of NIH-funded multisite trials produce demographically near-representative cohorts at every phase. Industry-led trials recover the established gradient (+1.95 pp). **The industry × academia joint trial shows the *steepest* race gradient of all: +4.65 pp from Phase I to Phase III, more than twice the slope of industry-solo, with Phase III African-derived enrollment in joint trials (8.2%) statistically identical to industry-solo (7.6%) and less than half of academia-led (19.0%) at the same trials’ own institutions**. When an academic medical center enrolls under an industry-written protocol, its catchment-population diversity is overwritten by the protocol’s eligibility criteria. The protocol launders the academic site’s home-population diversity out of the trial.

The **sex axis** carries a complementary signature on the same buckets. The pooled Phase I → Phase III female-share inversion documented above (41.4% → 48.7%) resolves almost entirely into one bucket: **industry-led trials, where female enrollment swings +10.0 percentage points from Phase I (39.6%) to Phase III (49.6%)**. Industry × academia joint trials carry a small residual inversion (+2.2 pp); academia-led and government-led trials are flat on the sex axis (Δ ≤ |0.7| pp). The industry-solo Phase III female-share is essentially at parity, but the Phase I share is not, and Phase I is where pharmacokinetic and pharmacodynamic baselines are calibrated. The same joint-trial vector that worsens the race gradient (laundering academic catchment-area diversity out) damps the female-share inversion that industry-solo trials show, the sex skew that industry-solo trials carry early is partly absorbed into the joint structure rather than amplified.

Both findings converge on the same structural locus. The site does not govern the demographics of the trial; the protocol does, and the protocol-writing function carries demographic loadings on both axes. Boards and credentialing bodies that govern protocol design, independent of the sponsor, are the structural intervention point on both the race and sex axes.

#### Reporting as a delayed governance signal

Race reporting itself, separately from enrollment, increased from 10.4% of trials pre-2017 to 59.6% post-2017 (χ^2^ = 15,996; *P* < 2.2 × 10^−16^; Mann-Kendall *Z* = 5.00, *P* = 5.7 × 10^−7^; Spearman ρ = 0.963, *P* = 1.6 × 10^−10^; **Fig. 4**). The inflection aligns with the 2017 NIH/OMB Revisions to the Common Rule mandate for race and ethnicity reporting^13^. But 40.4% of post-2017 trials still do not report race, eight years after an explicit regulatory instrument. The governance exists; the enforcement does not propagate.

**Fig. 4.**
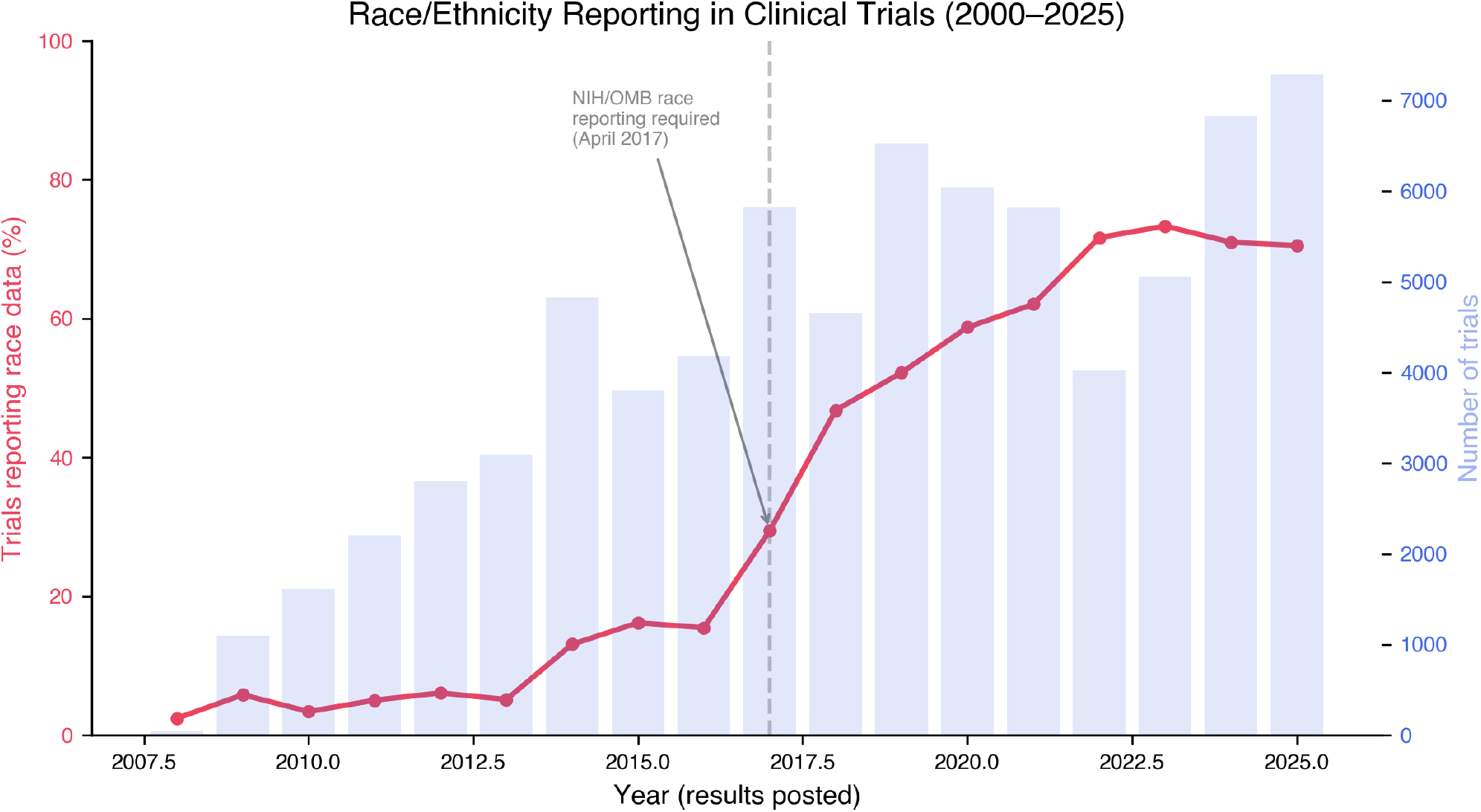
Race reporting over time (2008–2026) with the 2017 NIH/OMB mandate inflection. Proportion of trials reporting any race or ethnicity data, by year. Race reporting increased from 10.4% pre-2017 to 59.6% post-2017 (χ^2^ = 15,996; *P* < 2.2 × 10^−16^; Mann-Kendall *Z* = 5.00, *P* = 5.7 × 10^−7^; Spearman ρ = 0.963, *P* = 1.6 × 10^−10^). The inflection aligns with the 2017 NIH/OMB Revisions to the Common Rule mandate. Eight years after the mandate, 40.4% of trials still do not report race, the governance exists, the enforcement does not propagate.

The COVID-19 pandemic provided a natural experiment: did a global emergency disproportionately affecting African-derived communities^14,15^ change clinical trial diversity? It did not. COVID trials (10.8% African-derived, N = 3.98 million) were *less* diverse than non-COVID trials over the same window (18.1%, N = 21.64 million; Fisher’s OR = 0.55, *P* < 2.2 × 10^−16^). The massive vaccine trials (Pfizer, Moderna, BioNTech, 83.3% European-derived across 4 million participants) diluted diversity at scale. Industry barely moved: 6.7% pre-COVID to 8.3% post-COVID (+1.6 pp over five years, while publicly-funded research gained 6.1 pp).

### Sex axis and joint distribution

#### The two axes are independent failure modes

The phase-funnel and sponsor-class results above show that the same upstream-protocol bias signature, the CABG bedside template at population scale, is visible on race and sex separately. The natural next question, and the one the clinical-bias literature implicitly poses, is whether the two axes co-vary at the trial level: are the trials that exclude non-European-derived populations the same trials that exclude women?

We defined the **CABG-bias quadrant** as trials with `female_share < 0.50` AND `european_derived_share > 0.80` (male-skewed AND white-skewed) and tested its occupancy against independence of the two marginal distributions. Across the n = 37,737 eligibility-ALL trials with both sex and race data, **24.81% sit in the CABG-bias quadrant, against an expected 24.76% under independence** (χ^2^ = 0.13, *P* = 0.72) (**Fig. 6**). The Pearson correlation between female_share and european_derived_share across the same subset is r = 0.016. Per phase: the joint-quadrant occupancy is 26.2% Phase I, 33.2% Phase II, 22.8% Phase III, and 19.9% Phase IV, every phase indistinguishable from the rate independence predicts (all pairwise excess <1 pp, none surviving multiple-comparison correction).

The Phase III industry-vs-(academic+government) comparison on joint-quadrant occupancy is the only test that reaches significance (χ^2^ = 5.18, *P* = 0.023, Yates correction): academic execution is slightly more likely than industry execution to occupy the joint quadrant within Phase III (25.2% vs 22.3%), opposite to the race-only sponsor gradient. This inversion has the same explanation as the female-share gradient seen earlier: academic-led trials are predominantly US-domestic, and US-domestic enrollment runs whiter than the global enrollment industry trials reach. The Phase III academic+government bucket therefore concentrates white-skewed and male-skewed trials at slightly elevated rates, but the magnitude is small and reverses the inter-bucket race-axis ordering, a final piece of evidence that the two axes do not move together.

The naive reading of CABG would be that the same protocols which exclude women also exclude non-European-derived populations, that the bedside CABG bias generalizes as one reinforcing race × sex effect at the trial level. The data refutes that single-axis reinforcement reading. **CABG-style bias does not generalize as one effect with two readouts; it generalizes as one upstream cause with two independent readouts**. This is why we treat sex and race as co-equal demographic axes throughout: they require parallel architecture, not a unified composite. The race axis is quantified by the GEI immediately below; the sex axis is sketched as a parallel Sex Equity Index in the same section. Sex bias and race bias are co-equal failure modes of the same protocol-writing function, two parallel biases of the same upstream cause, and any structural intervention that closes one will not automatically close the other.

### Downstream: the genomic-database compounding

#### The Genomic Equity Index

The sparse matrix captures one layer of the failure. The database layer captures another. We combined both into a composite population-level metric.

For each population P, we computed seven subscores on a 0–1 scale: clinical trial enrollment ratio (this analysis), gnomAD coverage ratio^10^, GWAS Catalog ancestry share (computed from primary data, see Methods and GWAS Catalog Recomputation below), polygenic risk score transferability^12^, VUS resolution rate^16^, pharmacogenomic panel coverage^17^, and ClinVar submission volume^18^. The Genomic Equity Index weights these into a single scalar:

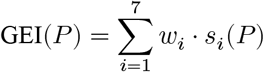

Weights (Methods) favor enrollment and polygenic accuracy (0.20 each) over reporting infrastructure (0.10 each) because the downstream clinical consequence flows from enrollment quality and predictive validity, not from counts of database entries. We assessed sensitivity by permuting the weights across 10,000 Monte Carlo iterations sampling from Dirichlet(1,1,1,1,1,1,1) with random seed 20260412 for deterministic reproducibility. European-derived populations were ranked first in 100% of iterations. The top-3 set (European-derived, Hispanic/Latino, Caribbean Admixed) was preserved in 75.3% of iterations; the bottom-3 set (Middle Eastern/North African, Indigenous American, Pacific Islander) was preserved in 86.9%; the full ranking achieved Kendall tau > 0.9 in 50.1% of iterations (mean tau = 0.910). Indigenous American and Pacific Islander populations occupied ranks 8 and 9 in 100% of iterations. The conclusion that European-derived populations are disproportionately advantaged and that Indigenous American and Pacific Islander populations occupy the equity floor is robust to the specific weight choice; ordinal positions among the middle-ranked populations are weight-sensitive and are reported with this caveat (**Extended Data Fig. 1**).

The results are in **Fig. 5** and **Table 3**. European-derived populations score 0.980. The remaining eight populations score 0.582 or lower. The total global equity gap, computed as (1−GEI(*P*)) × World Population(*P*) summed across populations, is **3.94 billion people receiving precision medicine calibrated for the 11% of humanity who are European-derived**.

**Table 3.** Genomic Equity Index by population (declared-weight ranking).

| Rank | Population | World % | GEI | Equity gap (millions) |
| --- | --- | --- | --- | --- |
| 1 | European-derived | 10.9 | 0.980 | 17.7 |
| 2 | Hispanic/Latino | 7.9 | 0.582 | 267.3 |
| 3 | East Asian | 21.5 | 0.443 | 969.2 |
| 4 | Caribbean Admixed | 0.3 | 0.434 | 13.7 |
| 5 | African / African-derived | 18.3 | 0.412 | 871.3 |
| 6 | Indigenous American | 0.5 | 0.332 | 27.1 |
| 7 | South Asian | 25.2 | 0.307 | 1,414.5 |
| 8 | Middle Eastern / North African | 5.2 | 0.239 | 320.5 |
| 9 | Oceanian / Pacific Islander | 0.6 | 0.227 | 37.6 |

**Fig. 5.**
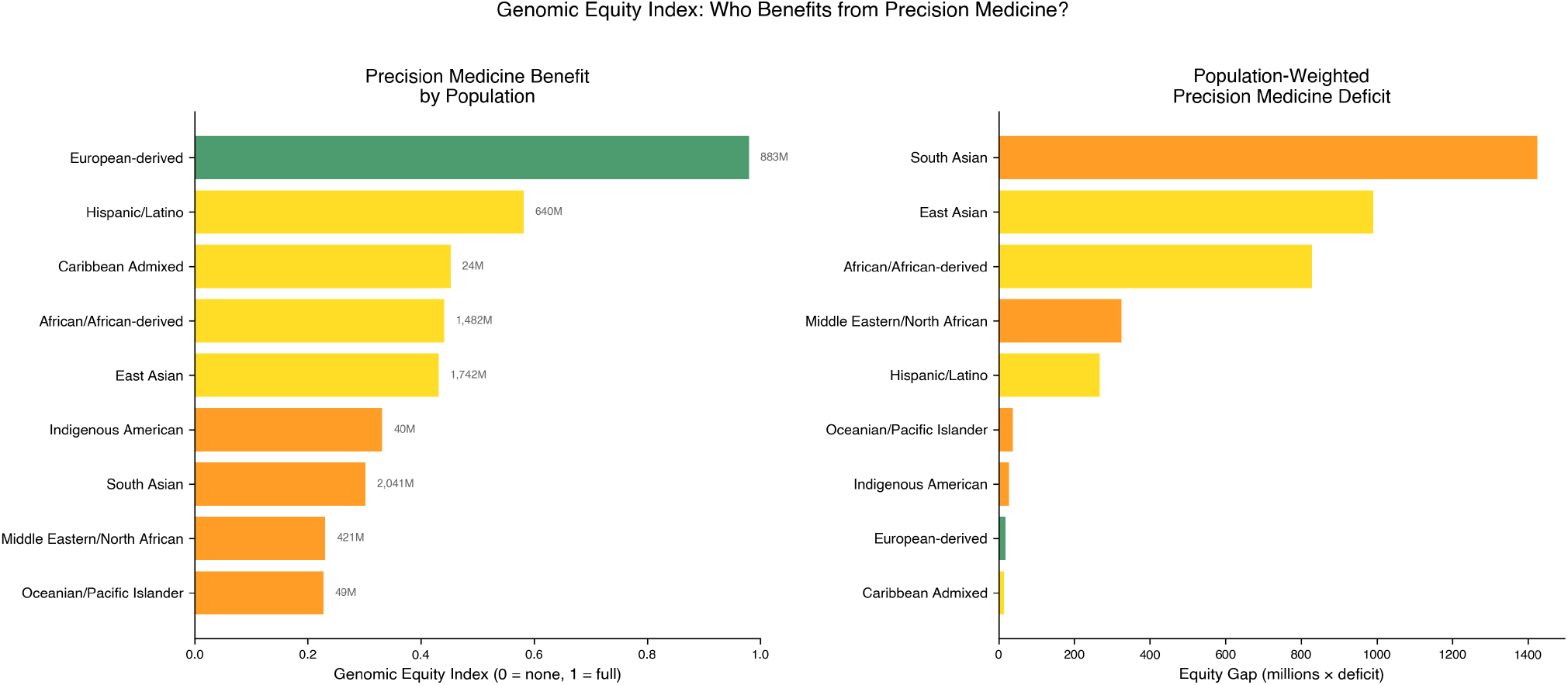
The Genomic Equity Index by population. Each bar is a population’s composite GEI on a 0–1 scale, weighted across seven subscores: clinical trial enrollment, gnomAD coverage, GWAS ancestry share, polygenic risk score transferability, VUS resolution rate, pharmacogenomic panel coverage, and ClinVar submission volume. European-derived populations score 0.980. Eight other populations score below 0.582. The right-axis annotation shows the global equity gap, the number of people receiving precision medicine calibrated for a population that is not theirs, totaling 3.94 billion across the eight underserved populations. The 11% of humanity that is European-derived has been the implicit reference population of the entire precision-medicine infrastructure.

South Asian populations (2.04 billion, GEI 0.307) carry the largest absolute gap: 1,414 million people-deficit units. The precision medicine literature has concentrated on African-derived and Latino disparities; the GEI reveals that South Asian populations, India, Pakistan, Bangladesh, Sri Lanka, face the largest absolute deficit and among the smallest per-capita allocation of precision medicine infrastructure. The global equity gap summed across populations is **3**,**939 million people-deficit units**, 3.94 billion people receiving precision medicine calibrated for the 11% who are European-derived.

#### A parallel Sex Equity Index

The GEI is the dosimeter for the race-axis manifestation of the upstream-protocol bias. The same seven-subscore architecture is portable to the sex axis as a Sex Equity Index (SEI), and we have computed two of the seven subscores in this paper: trial enrollment (the female_share gradient documented above) and GWAS participation (sex stratification is recoverable from the same NHGRI-EBI primary download). The remaining five subscores, gnomAD coverage by sex, polygenic risk score transferability by sex, VUS resolution by sex, pharmacogenomic panel coverage by sex, and ClinVar submission volume by sex, are infrastructure tasks not solved here, because the underlying genomic resources do not currently surface sex stratification in the same way they surface ancestry stratification. This asymmetry, sex compliance high at the trial layer (99.3%) but low or absent at the genomic-database layer, is itself a finding: the data shadow on the sex axis flips polarity between the upstream (CT.gov) and downstream (gnomAD/ClinVar/PRS) infrastructure. We flag the SEI as the natural next-paper construct, computable on the same architecture once the genomic-database layers expose sex stratification at parity with ancestry.

#### Caribbean evidence: ancestry-specific drug response in the same populations the matrix lists in black

The Caribbean Admixed population sits at GEI rank 4 (gap 13.7M), but the deficit is the per-capita extreme: this is the population for which Sirugo, Williams and Tishkoff^5^ documented “missing diversity” most acutely, and for which the trial pipeline returns ZERO oncology evidence in the sparse matrix above. Direct evidence that the resulting drug-evidence gap matters at the population level comes from the Caribbean-anchored clinical research literature, much of it co-authored by members of the present byline. Haraksingh and colleagues recently demonstrated **ancestry-specific drug vulnerabilities in an Afro-Caribbean prostate cancer model with therapeutic implications for Black patients**^19^, and **CYP2C19*2 /* 3 allelic variants predicting altered clopidogrel response in a Trinidad & Tobago cardiovascular cohort**^20^ (the same population whose recurrent-stroke and cardiovascular outcomes the precision-medicine-credentialing thesis depends on). On the cancer axis specifically, **ancestry-specific BRCA1-185delAG screening of formalin-fixed Trinidad & Tobago breast cancer biopsies**^21^ documents an actionable founder mutation in the same Caribbean Admixed population whose oncology cell of the sparse matrix is empty, paired with **a 2015 multi-cohort analysis of ancestry, geography, and breast cancer incidence, mortality, and survival in Trinidad & Tobago**^22^ that established the demographic outcome substrate the present trial-pipeline analysis is silent on. On the LMIC clinical-trial-methodology axis, **a 2025 cohort study of HIV durable viral suppression among clinic attendees in Trinidad & Tobago**^23^ shows that the same population can be investigated to publication-grade methodological standards when researchers choose to investigate; the absence in the trial pipeline is not an absence of investigability, but an absence of investigation. All four findings are consistent predictions of the sparse-matrix and LD-paradox architecture: the populations the matrix lists in black are precisely the populations in which ancestry-specific drug response and disease structure exist and are detectable when investigated, and the empty cells of the matrix are the absence of investigation, not the absence of biology.

#### The linkage disequilibrium paradox

The economic irrationality of the extraction pattern is sharpest at the biological level. African-derived populations carry shorter linkage disequilibrium (LD) blocks than European-derived populations^24^, the signature of 300,000 years of ancestral recombination in Africa versus approximately 70,000 years of bottlenecked recombination outside it. This cuts both ways. Shorter LD means GWAS tag-SNP arrays designed around European LD structure (where long blocks allow one SNP to tag many neighbours) perform worse in African-derived genomes^1,12^: the tag cannot carry the signal when the block is too short. African-derived studies therefore demand larger sample sizes to achieve the same statistical power. The cost is real.

But shorter LD also yields *finer mapping resolution*. In European-derived genomes, a GWAS hit lands in a long LD block, hundreds of kilobases of correlated variants, any one of which could be causal. In African-derived genomes the same locus is dissected into shorter blocks. The causal variant is easier to isolate. The drug target is sharper. PCSK9 is the proof^25^: the loss-of-function variants (Y142X, C679X) that launched a $3 billion annual drug class were discovered in African-derived participants in the Dallas Heart Study, not because African-derived populations were studied more but because the LD architecture permitted the dissection that European LD blocks were too blunt to resolve. The gain-of-function variants were identified first in French families^26^, the fluke. The loss-of-function variants, the drug target, required the population whose genome architecture provides the highest resolution.

The population whose genome architecture would produce the best drug targets is the population the pharmaceutical industry invests least in studying. The population that demands larger N (more expensive) is the population that delivers finer resolution (more valuable). The sparse matrix captures the economic irrationality at the industrial level. The LD paradox captures it at the biological level.

## Discussion

### The upstream-bias thesis is confirmed on both demographic axes

We took the named bedside phenomenon CABG^9^, a sex disparity that originates in diagnostic timing rather than at the operating-room door, and asked whether the same upstream-bias model is visible at the population scale of clinical-trial enrollment. The answer is yes, and on both axes simultaneously. On the race axis, African-derived enrollment declines monotonically Phase I → Phase III in industry-led trials (14.3% → 8.1%; Δ Phase I → III = −3.1 pp/phase), Indigenous American and Pacific Islander populations occupy evidence deserts in 15 of 15 therapeutic areas, and industry oncology Phase III reaches 1.8% African-derived against a 15% mortality burden. On the sex axis, female enrollment runs the opposite direction (41.4% Phase I → 48.7% Phase III in the eligibility-ALL subset), with the inversion concentrated in industry-solo trials (+10.0 pp). Reporting compliance is asymmetric (99.3% sex vs 61.6% race) but in both cases the bias signature visible post hoc in the recorded enrollment is a signature whose locus is upstream of the recorded efficacy result, exactly as the CABG bedside model predicts.

### Independence is the load-bearing structural finding

The clinical-bias literature’s natural expectation is that the same protocols that exclude non-European-derived populations also exclude women, that the bedside CABG phenomenon generalizes as one race × sex reinforcing effect at the trial level. The data refutes that. Across 37,737 eligibility-ALL trials, the joint CABG-bias quadrant occupancy is 24.81%, exactly the rate predicted by independence of the two marginals (24.76%; χ^2^ = 0.13, *P* = 0.72). Pearson r = 0.016. Per phase, every joint-quadrant test is indistinguishable from independence. **CABG-style upstream bias does not generalize as one effect with two readouts; it generalizes as one upstream cause with two independent readouts**. Any structural intervention that closes one will not automatically close the other, and the equity argument requires parallel architecture rather than a unified composite, which has direct consequences for policy design and for measurement.

### Quantification: GEI today, SEI tomorrow

The Genomic Equity Index is the dosimeter of the race-axis manifestation of the upstream-protocol bias. Its 7-subscore architecture, clinical-trial enrollment, gnomAD coverage, GWAS Catalog ancestry share (recomputed from primary data; 88.3% European-derived, not 78% as published^11^), polygenic risk score transferability, VUS resolution rate, pharmacogenomic panel coverage, and ClinVar submission volume, yields a global equity gap of 3.94 billion people receiving precision medicine calibrated for the 11% who are European-derived; European-derived populations score 0.980 and eight others below 0.582; the top and bottom rankings are robust in 100% of 10,000 Dirichlet weight permutations. The same architecture is portable to a Sex Equity Index: trial enrollment and GWAS participation are computable for sex now (the SEI for those subscores resolves to a parallel scalar today), but the remaining five subscores require the genomic-database layer to surface sex stratification at parity with ancestry, which it currently does not. The data shadow on the sex axis is high at the upstream trial layer (99.3% reporting) and low at the downstream genomic-database layer, flipped polarity from the race axis. The SEI is the natural next-paper construct on the same architecture; the GEI is the bread-and-butter quantification this paper delivers.

### Governance closure on both axes

The empty cells of the sparse matrix and the early-phase female-deficit are not evidence of absence; they are evidence of governance, of which populations were counted, in which phase, by whom. That governance was made cell by cell, protocol by protocol, by pharmaceutical sponsors and their academic execution partners whose trials are registered in a public database. The correction is governance too. **The locus is the protocol-writing function**: industry-led trials produce the race extraction gradient and the sex inversion; the industry × academia joint trial worsens the race gradient (laundering academic catchment-area diversity out) while damping the sex inversion; academia-led and government-led trials remain near-flat on both axes. **Boards and credentialing bodies that govern protocol design, independent of the sponsor, are the structural intervention point on both axes**. The senior author’s pre-2015 publication record exemplifies the pattern from inside it (PennCNV^27^, the autism CNV studies^28,29^, the age-related macular degeneration GWAS series^30,31^, the consortium-scale GWAS work on childhood obesity / intracranial volume / infant head circumference / pubertal timing^32–35^, and the STARGEO open-data infrastructure^36^, all in discovery cohorts that were 90–100% European-derived); documenting the pattern from inside it is a precondition for correcting it. The FDA Diversity Action Plan guidance^37^, removed in January 2025, partially restored in February 2025, is one regulatory instrument; it addresses race but not sex, and it addresses recruitment targets but not protocol-writing governance. The data remains.

Precision medicine is precise. It is just not precise for most of humanity. The correction does not require a new biobank; it requires governing the upstream protocol-writing step on both axes, on every trial, before the bias becomes a label.

## Methods

### Data acquisition

ClinicalTrials.gov API v2 was queried on 1 May 2026 with cursor-based pagination (pageSize = 1000) and filter `AREA[HasResults]true`, returning 77,770 studies with posted results across all conditions, phases, and statuses. No studies were excluded. The complete download required 78 API requests.

### Race extraction

Baseline race and ethnicity were extracted from `resultsSection.baselineCharacteristicsModul e`, normalized to the NIH/OMB standard (White, Black or African American, Asian, AI/AN, NH/PI, More Than One Race, Other, Unknown). Per-trial `european_derived_share = White / (Sum of known-race categories)`, dropping “Unknown or Not Reported”.

### Sex extraction

Baseline sex was extracted from the same baseline-characteristics module. Per-trial `female_share = F emale / (Female + Male)`. Protocol-level `eligibilityModule.sex` (ALL / FEMALE / MALE) was additionally extracted so sex-restricted protocols (breast cancer, prostate cancer, OB/GYN, etc.) could be distinguished from discretionary sex skew in eligibility-ALL trials. 9,333 sex-restricted protocols (6,358 female-only, 2,975 male-only) were filtered, leaving n = 37,737 eligibility-ALL trials reporting both sex and race as the joint analytic subset.

### Therapeutic area assignment

Each trial’s primary condition was mapped to one of 15 MeSH-derived therapeutic areas (Extended Data Table 1). Trials with no MeSH mapping were excluded from the therapeutic-area analysis.

### Evidence Score, sparse matrix, GEI

Definitions in main text; classification ADEQUATE (ES ≥ 0.30), WEAK (0.10–0.30), INSUFFICIENT (0.01–0.10), ZERO (<0.01). GEI subscores: clinical trial enrollment (this analysis), gnomAD coverage (Karczewski et al.^10^, v4), GWAS Catalog ancestry share (primary-data recomputation), PRS transferability (Martin et al.^12^), VUS resolution (Caswell-Jin et al.^16^), PGx coverage (Gaedigk et al.^17^), ClinVar submissions (Landrum et al.^18^). Weights: trial 0.20, gnomAD 0.15, GWAS 0.15, PRS 0.20, VUS 0.10, PGx 0.10, ClinVar 0.10.

### GWAS Catalog recomputation

`gwas-catalog-ancestry.tsv` was downloaded from the NHGRI-EBI FTP release on 12 April 2026 (279,343 rows; SHA-256 attested in `data/gwas_catalog/PROVENANCE.json`). Rows were parsed, individuals summed per broad ancestral category, multi-ancestry rows split evenly, and mapped onto the 9-population GEI framework. The `African American or Afro-Caribbean` label was split 70/30 between African-derived and Caribbean Admixed (documented in `PROVENANCE.json`; does not affect top or bottom GEI ranks under sensitivity). Primary-data recomputation yielded **88.3%** European-derived participation across 13.5 billion cataloged participants, higher than the 78% in Mills & Rahal 2020^11^. This recomputation is the sole authoritative GWAS subscore source.

### Joint sex × race analysis

The CABG-bias quadrant is defined as `female_share < 0.50` AND `european_derived_ share > 0.80` (male-skewed AND white-skewed), the named bias quadrant from the clinical CABG literature. For each phase × sponsor-class cell observed quadrant occupancy was computed against the rate predicted under independence of the two marginals (P(female<0.50) × P(ED>0.80)), tested with a Yates-corrected χ^2^ 2×2 contingency. Pearson r between `fema le_share` and `european_derived_share` was computed across the n = 37,737 analytic subset. The Phase III industry-vs-(academic+government) comparison on quadrant occupancy was tested with the same Yates-corrected χ^2^. Computation by `analyze_sex_race_stratification.py` and figure by `figure_sex_race_stratification.py`, with SHA-256 attestation in `data/sex_race_PROVENANCE.json`.

### Sensitivity analysis

GEI weights permuted across 10,000 Monte Carlo iterations from Dirichlet(1,1,1,1,1,1,1) at random seed 20260412 for deterministic reproducibility. Per-iteration ranking distributions in `data/gei_sensitivity.json` (computed by `sensitivity_gei.py`).

### Statistical tests

Chi-squared goodness-of-fit (Census 2020 reference); Clopper-Pearson exact 95% CIs; Mann-Kendall for temporal signals; Fisher’s exact for phase and sponsor comparisons; Bonferroni correction at α = 0.003 for 15 therapeutic areas. scipy 1.17.1, Python 3.14.

### Reproducibility

Every figure and table can be regenerated from source.

## Extended Data

**Extended Data Figure 1.**
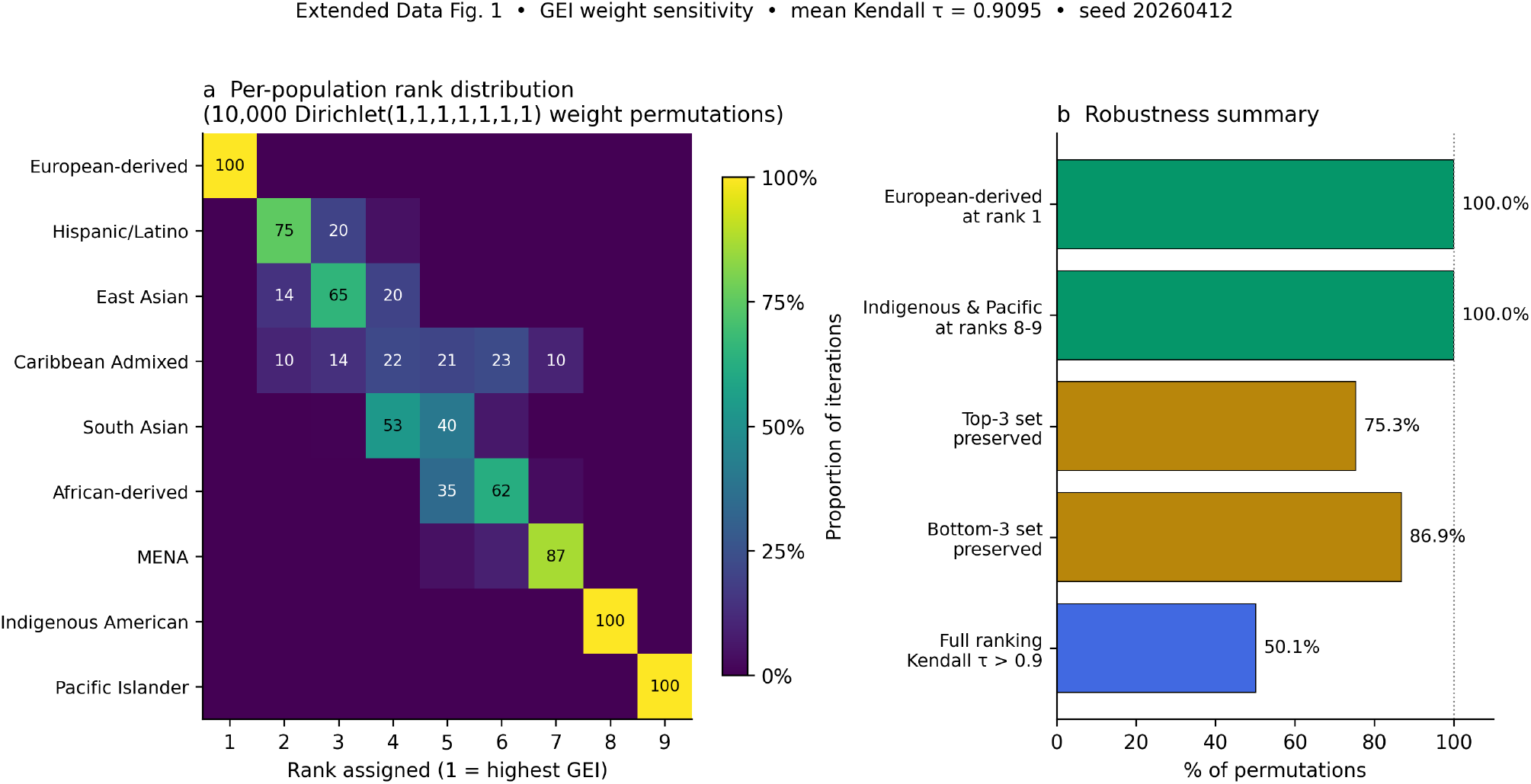
GEI sensitivity analysis. Population rankings across 10,000 Monte Carlo iterations sampling weights from Dirichlet(1,1,1,1,1,1,1) with random seed 20260412. European-derived populations ranked first in 100% of iterations. The top-3 set (European-derived, Hispanic/Latino, Caribbean Admixed) was preserved in 75.3% of iterations; the bottom-3 set (Middle Eastern/North African, Indigenous American, Pacific Islander) was preserved in 86.9%. Indigenous American and Pacific Islander populations occupied ranks 8 and 9 in 100% of iterations. Full ranking Kendall tau > 0.9 in 50.1% of iterations (mean tau = 0.910). The conclusion is robust to weight choice for top and bottom positions; ordinal positions among middle-ranked populations are weight-sensitive.

**Extended Data Figure 2.**
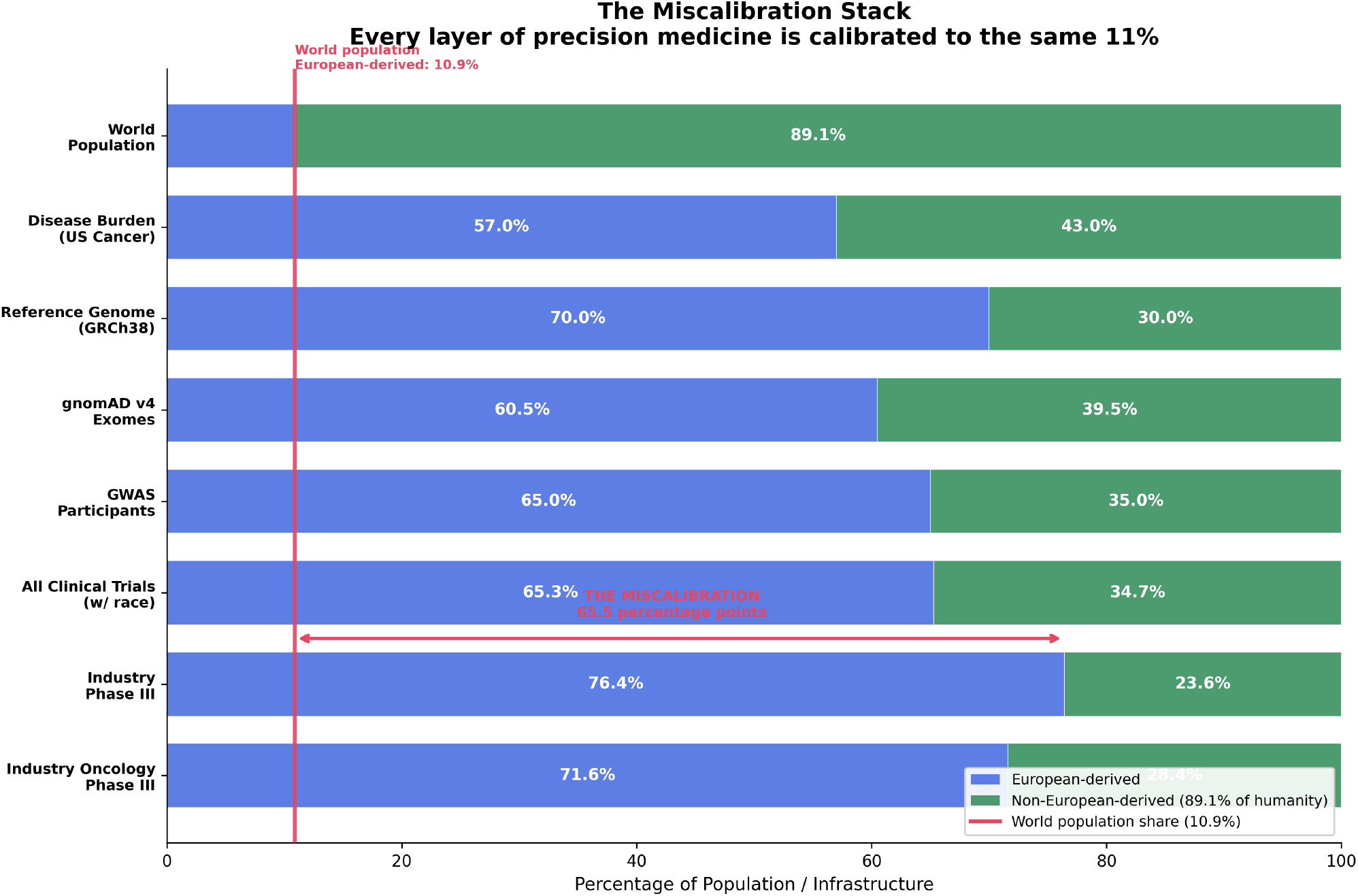
The miscalibration stack across the precision-medicine pipeline. Layered comparison of European-derived (blue) and non-European-derived (green) representation across the 8 stages of the precision-medicine pipeline: world population (10.9% European), US cancer disease burden (57%), GRCh38 reference genome (70%), gnomAD v4 exomes (60.5%), GWAS Catalog participants (65%), all clinical trials reporting race (65.3%), industry Phase III trials (76.4%), industry oncology Phase III (71.6%). Red vertical line marks the world-population European-derived share (10.9%). The compression begins as a moderate sampling bias at the genome reference layer and amplifies through every downstream stage, ending with industry oncology Phase III at ∼7× the world share.

## Author Contributions

**Roles and affiliations. Anil Bajnath, MD, MBA** is Founder & President of the American Board of Precision Medicine and a Director of the CANONIC Foundation. **Allana Roach, PhD, MSB** is the CANONIC Ethics Board Lead and a Senior Associate Scientist at the American Cancer Society, Extramural Discovery Science. **Rajini Haraksingh, PhD** is Lecturer in Biotechnology at The University of the West Indies, St. Augustine, and Director of Panomics & Chair of the Biobanking Working Group at the Yale-TCC Consortium (NIH/NIMHD-funded precision medicine and minority health disparities; Caribbean / UWI institutional anchor). **Irman Forghani, MD, FACMG** is Director of Clinical Genetics at Mount Sinai Medical Center of Florida, FACMG-certified clinical molecular geneticist. **Alexander N. Evans, MD, MBA, FACS** is Associate Professor of Surgery at Howard University College of Medicine, Trauma Medical Director at Howard University Hospital, and Co-Director of the Howard AI in Healthcare Consortium (HBCU clinical-AI lead). **Elena Cyrus, PhD, MPH** is the CANONIC Chief Advisor for Global Health and Associate Professor of Medicine at the University of Central Florida College of Medicine; former Phase II/III HIV-prevention trial PI / protocol manager (HPTN, MTN, FHI360, Yale-Fogarty Lima). **Dexter Hadley, MD, PhD** is Founder and Chair of the CANONIC Foundation and Director of AI at the American Board of Precision Medicine.

### Contributions to the manuscript (CRediT taxonomy)

- **A.B**. — Conceptualization; Funding acquisition (ABOPM); Project administration; Writing — review & editing. Anchored the precision-medicine governance frame and the ABOPM credentialing context that motivated the upstream-bias hypothesis at population scale.
- **A.R**. — Conceptualization; Validation; Writing — review & editing. Provided the equity and community-health framing and the ACS Extramural Discovery Science perspective on how trial demographics propagate into population-level evidence.
- **R.H**. — Methodology; Validation; Writing — review & editing. Contributed the panomics and linkage-disequilibrium architecture, the Caribbean institutional anchor, and the genomic-database layer of the GEI calibration.
- **I.F**. — Conceptualization; Methodology; Investigation; Writing — review & editing. Drove the sex × race × phase × sponsor stratification (Fig. 6) and the joint CABG-bias quadrant analysis after surfacing the sex axis as a missing demographic dimension.
- **A.N.E**. — Conceptualization; Validation; Writing — review & editing. Provided the surgical and acute-care decision context (Howard AI in Healthcare Consortium) and the HBCU institutional perspective on protocol-level bias.
- **E.C**. — Conceptualization; Methodology; Validation; Writing — review & editing. Anchored the HIV-prevention Phase II/III protocol-design background (HPTN, MTN, FHI360, Yale-Fogarty Lima) and the global-health framing of the Genomic Equity Index.
- **D.H**. — Conceptualization; Methodology; Software; Formal analysis; Investigation; Data curation; Visualization; Writing — original draft; Writing — review & editing; Supervision; Project administration. Designed the Clinical-Trials.gov v2 audit pipeline, constructed the Genomic Equity Index, performed the sponsor-class gradient analysis, recomputed GWAS-Catalog ancestry from primary data, and authored the manuscript.

**Fig. 6.**
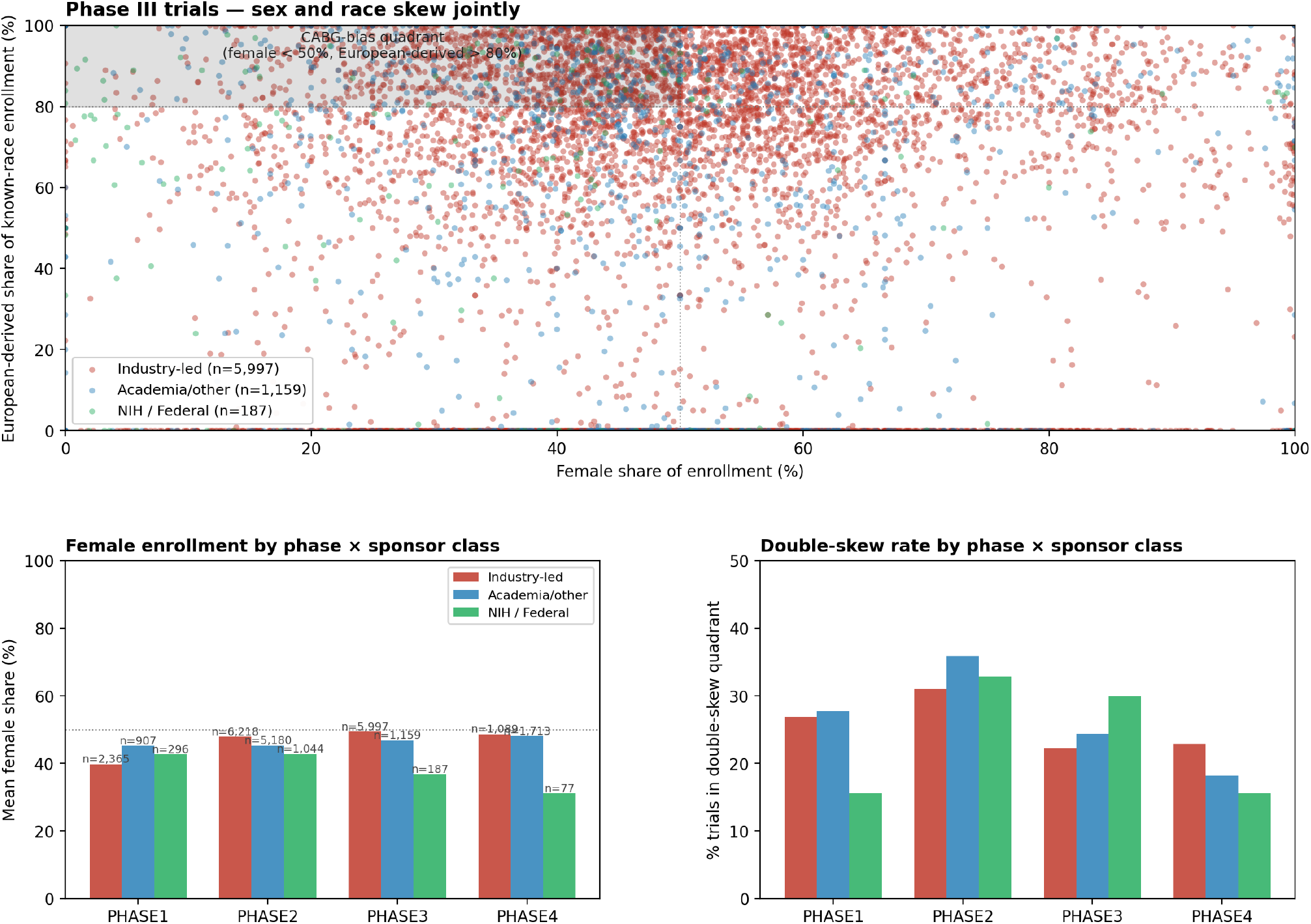
The two axes are statistically independent. (a) Phase III trials, female_share vs european_derived_share scatter, colored by sponsor class; n = 5,997 industry-led, 1,159 academia/other, 187 NIH/Federal. The CABG-bias quadrant (female_share < 0.50 AND european_derived_share > 0.80) is shaded. (b) Female enrollment by phase × sponsor class, academic trials run ∼5 percentage points higher female-share than industry across every phase. (c) CABG-bias quadrant occupancy by phase × sponsor class. Pooled across 37,737 trials, observed quadrant rate (24.81%) is statistically indistinguishable from the rate predicted by independence of the two marginals (24.76%; χ^2^ = 0.13, *P* = 0.72). Pearson r = 0.016. The Phase III industry-vs-academic comparison reaches χ^2^ = 5.18, *P* = 0.023, with academic execution slightly more likely than industry to occupy the joint quadrant. Sex skew and race skew co-occur in the joint quadrant only because their marginals coincide there.

## Acknowledgments

The authors thank the ClinicalTrials.gov v2 API maintainers (NLM/NIH) for sustained public access to the trial-results corpus, the NHGRI-EBI GWAS Catalog team for primary-data access, and the seven coauthors’ home institutions for protected research time during manuscript preparation.

## Data Availability

The complete dataset (77,770 studies; data freeze 1 May 2026), the NHGRI-EBI GWAS Catalog primary download (with SHA-256 in `data/gwas_catalog/PROVENANCE.json`), the collaborator/responsible-party enrichment (`trials_collab orators.jsonl`, also SHA-256 attested), the 13 analysis scripts (`download_trials.py`, `download_collaborators.p y`, `analyze_race_enrollment.py`, `analyze_sparse_matrix.py`, `analyze_genomic_equity.py`, `analyze_gwas_ ancestry.py`, `analyze_sponsor_class_gradient.py`, `analyze_sex_race_stratification.py`, `forensic_pha rma.py`, `sensitivity_gei.py`, statistical-test, figure, and sponsor-gradient figure scripts), and all intermediate data files are at the project’s public source repository (CLINICAL-TRIALS-RACE manuscript directory). The dataset and scripts are cryptographically attested with a SHA-256 hash chain; the content hash of this manuscript at submission is recorded in the CANONIC governance ledger via `MANUSCRIPT_CONTENT_MINTED` and `MANUSCRIPT_EXPORT_SNAPSHOT` events per `the CANONIC governance LEDGER service contract § Event Types`.

